# COMPARING EFFICACY AND SAFETY OF LEFT ATRIAL APPENDAGE CLOSURE DEVICES: A NETWORK META-ANALYSIS OF RANDOMIZED CONTROLLED TRIALS

**DOI:** 10.1101/2024.03.18.24304489

**Authors:** John W Davis, Steven L Mai, Wissam Harmouch, Jenna Reisler, Micaela MacKay, Elizabeth Davis, Mike Rich

## Abstract

**Introduction:** Atrial fibrillation-related stroke is a leading cause of morbidity and mortality. Options for prevention include left atrial appendage closure devices or oral anticoagulation. However, it remains unclear which option may be superior overall.

**Methods:** We conducted a systematic review and network meta-analysis of all clinical trials comparing the WATCHMAN, Amplatzer Amulet (Amulet), and/or OACs. The primary outcomes of interest were any stroke and all-cause death. Safety outcomes included any thromboembolism, device embolization, and pericardial effusion. We calculated risk ratios and heterogeneity statistics for each comparison, and calculated the probability of intervention superiority where at least one comparison was significant.

**Results:** There were 441 articles identified from the search, from which 5 eligible RCTs were identified (n=1,811). Compared to OACs (all warfarin), risk of stroke was non-significantly decreased with WATCHMAN (RR=0.90, 95% CI: 0.48, 1.72, I^2^=13.4%), but risk with Amulet was non-significantly lower than WATCHMAN (RR=0.95, 95% CI: 0.50, 1.81). However, observed risk of all-cause death was significantly lower with Amulet than OAC (RR=0.53, 95% CI: 0.33, 0.85, I^2^=0%) and trended towards significance versus WATCHMAN (RR=0.74, 95% CI: 0.55, 1.02, p=0.06). The P-score was 0.982, signifying a >98% probability Amulet was superior to all alternatives. Risk of thromboembolism was non-significantly increased with WATCHMAN (RR=2.04, 95% CI: 0.23, 18.4) and Amulet (RR=1.54, 95% CI: 0.11, 22.1), with head-to-head comparison favoring Amulet (RR=0.76, 95% CI: 0.17, 3.38, I^2^=0%). Versus WATCHMAN, device embolization risk was non-significantly elevated with Amulet (RR= 2.38, 95% CI: 0.67, 8.43, I^2^=0%). Finally, risk of pericardial effusion was significantly elevated with Amulet versus OACs (RR=27.0, 95% CI: 3.48, 210) and versus WATCHMAN (RR=2.11, 95% CI: 1.41, 3.17, I^2^=0%). The inverse P-score for Amulet (0.9995) indicated a very high probability Amulet was inferior to alternatives.

**Conclusion:** While risk of some adverse events was greater with Amulet, we estimated >98% probability Amulet is superior to alternatives in risk of death. Pooled patient-level analyses are warranted.

## Introduction

Despite advances in management, stroke is the second leading cause of death and the third leading cause of disability worldwide.^1^ Atrial fibrillation (AF) is the most common underlying cause of cardioembolic stroke and increases an individual’s risk of stroke 5-fold.^2^ The left atrial appendage (LAA) is a common source of emboli which has been implicated in approximately 90% of AF-related stroke.^2^ Compared to strokes secondary to carotid disease, strokes related to LAA thrombi are associated with greater disability burden and higher mortality overall.^3^

Although oral anticoagulation effectively reduces risk of stroke and thromboembolism among patients with AF,^4^ these agents require medication adherence and are associated with bleeding complications which may be fatal.^5^ Thus, percutaneous left atrial appendage closure (LAAC) devices have been developed to prevent thrombus embolization and cardioembolic stroke in these patients.^6^ The Watchman device, a single-seal LAAC device approved by the FDA in 2015, prevents thromboembolic events with efficacy noninferior to warfarin.^6^ The Amplatzer Amulet (Amulet) device was approved for LAAC in 2021 after demonstrating non-inferiority in preventing ischemic stroke and systemic embolism when compared to Watchman.^6^ Compared to Watchman, Amulet deploys multiple discs to seal the LAA, which allows for periprocedural adjustment and possibly greater deployment success rates.

Primary safety analysis from the Amulet IDE demonstrated higher rates of procedural-related complications, such as pericardial effusions and device embolization, when compared to Watchman.^6^ However, some important events such as all-cause mortality were underpowered for analysis in these trials. A network meta-analysis utilizes both direct and indirect estimations from included studies to increase power for detecting small but important associations. The present network meta-analysis (NMA) aims to investigate the comparative safety and efficacy of LAAC devices, both head-to-head and against oral anti-coagulation (OAC).

## Methods

### The trials

PubMed, Cochrane Database, and Web of Science were searched for systematic reviews, meta-analyses and/or randomized clinical trials published in English on Watchman and/or Amplatzer (Amulet) devices. The search dates were from 1946 to May 2023. Methods were registered prospectively with Prospero (CRD# 42023428226; see supplement for search terms and strategy). We aimed to include all RCTs comparing either device to anticoagulation (either warfarin or direct oral anticoagulants, DOACs), or to the alternative device for subjects with AF. There were no additional exclusion criteria for trial eligibility. At least 2 authors blinded to codification by other reviewers identified trials for final inclusion, then coded each for quality with a five-point Jadad quality score.^7^ Data on exposure, outcomes, comorbidities, and trial design were also extracted and collated for analysis. We also searched clinicaltrials.gov for any additional information, and reported whichever source was more comprehensive for each outcome. After independent extraction, any disagreements were arbitrated by the principal investigator (JD). We made attempts to contact trial investigators for discrepancies.

### Exposure variable

Study arms were classified as ‘Watchman-exposed’, ‘Amulet-exposed’, or ‘Oral Anti-Coagulation’ (OAC)-exposed.

### Outcome variables

We first aimed to code for efficacy of the intervention – prevention of stroke (either ischemic, embolic, or hemorrhagic) and/or death. Adjudication as intervention-related or unrelated was ignored for all outcomes to avoid bias and because of variable definitions among trials. Trial definitions for each outcome were used because these data are study-level and cannot be reliably abstracted otherwise.

The first outcome was for any stroke (ischemic or hemorrhagic). The second was death from any cause. Next, we assessed adverse event outcomes. The first adverse event codified was risk of any thromboembolism. Next, we coded for risk of device embolization. Finally, we assessed risk of pericardial effusion, a known complication of the procedure.^8,9^ All events were counted regardless of whether trialists identified the event as ‘intervention-related’.

### Analysis

We largely followed methods from previous analyses^10^ in conducting this study. Published aggregate data from each trial were used. Prevalence estimates were calculated for each outcome by intervention arm. Pooled relative risk (RR) was estimated for each outcome, as follow-up time was approximately equal across each intra-study arm. Given that some events, such as device embolization, are obviously impossible with OAC controls, we used a 0.5 continuity correction to generate RRs where 0 cells were present.

When conducting the network meta-analysis (NMA), we utilized the Inverse Variance method with random effects in the ‘netmeta’ package^11^ to generate risk ratios for each outcome. We assessed the percentage of evidence utilized for each comparison that is ‘indirect’ (comparing interventions across trials) for each outcome. Risk of bias related to heterogeneity was assessed for all trials using standard measures (I^2^, Cochran’s Q)^12,13^ in addition to generating funnel plots. High risk of heterogeneity bias was defined as >75% I^2^.^12,13^ Because there was a small number of eligible trials, meta-regression and formal hypothesis tests for bias were not feasible. We qualitatively assessed the transitivity assumption – that there is no effect modification with baseline characteristics of subjects across trials – by confirming that age, comorbidities, and AF-related stroke risk were approximately equal at baseline. League tables providing RR estimates of all available comparisons were generated to display multiple comparisons.

Because NMA indirect evidence breaks randomization, causal language may not be appropriate.^14^ We therefore avoid doing so here. However, we used a frequentist tool (‘netrank’ command in the netmeta package)^11,15^ to quantify the probability that one intervention was superior to alternatives when at least one comparison was significantly different. A ‘P-score’, the inverse of a one-sided hypothesis test of superiority, was estimated for each outcome. A P-score of < 0.05 suggests there is less than a 5% probability that the intervention is superior to alternatives, whereas a P-score > 0.95 indicates a 95% probability the intervention is superior to alternatives.^15^ For ease of interpretation, we report inverse P-score (the probability of inferiority) where one intervention appears worse than alternatives.

We followed PRISMA guidelines for reporting and analyzing these studies. All analyses were conducted in R, Version 4.1.3.^16^

## Results

### Search Results

There were 441 articles that were initially returned in our search, of which 33 (7.5%) were either RCTs, systematic reviews, or meta-analysis that contained potentially eligible trials. From these, we identified 5 RCTs^8,9,17–19^ that met criteria and were abstracted for data. Three^9,18,19^ assessed both LAAC devices head-to-head, whereas the other 2 trials^8,17^ assessed WATCHMAN to OACs (both warfarin). Because we were interested in comparative efficacy of devices, trials that aggregated reports of WATCHMAN and Amulet (i.e., PRAGUE-17)^20^ versus OACs could not be included. There were no identified trials comparing Amulet to OACs. The PRISMA diagram (**Figure 1**) displays the search study flow.

### Study Quality and Subjects Enrolled

Summaries of study quality and characteristics are found in **Table 1**. Jadad scores for study quality varied. Because subjects and healthcare providers could not be blinded to WATCHMAN vs. OAC, the 2 earliest trials^8,17^ scored 3/5 for study quality. Two^9,18^ additional trials scored 4/5 possible points (Lakkireddy,^9^ lack of echo tech blinding; Mansour,^18^ time sequence-based randomization), and 1 trial^19^ scored 5/5 possible points for quality. Of note, the 2 trials assessing WATCHMAN vs. OAC excluded subjects with contraindications to warfarin, whereas the 3 trials assessing LAAC devices head-to-head did not. There were no other obvious differences in baseline comorbidities across interventions.

**Table 1:** Descriptive Statistics.

| Table 1: Descriptive Statistics |  |  |  |  |  |  |  |  |  |  |
| --- | --- | --- | --- | --- | --- | --- | --- | --- | --- | --- |
| Author, Year (Trial Name) | Comparison groups | N (Total ) | N (Watchman) | N (Amulet/Warfarin) | Watchman Gen. | Exc. if Warfarin CI | Age (Mean) | Female( %) | Mean F/u (d) | Jad ad |
| Galea 2022 (SWISS-APERO) | Watchman vs Amulet | 221 | 110 | 111 | FLX | Y | 76.9 | 29.4 | 45 | 5 |
| Lakireddy 2021 (Amulet IDE) | Watchman vs Amulet | 1878 | 944 | 934 | 1st Gen. | N* | 75 | 40 | 540 | 4 |
| Mansour 2021 | Watchman vs Amulet | 51 | 25 | 26 | 2.5/FLX | N | 75.5 | 23.5 | 365 | 4 |
| Holmes 2014 (PREVAIL) | Watchman vs Warfarin | 407 | 269 | 138 | 1st Gen. | Y | 74.3 | 30 | 540 | 3 |
| Holmes 2009 (PROTECT AF) | Watchman vs Warfarin | 707 | 463 | 244 | 1st Gen. | Y | 72 | 29.7 | 540 | 3 |
| *Long term OAC Contraindication, but considered safe for short-term warfarin therapy |  |  |  |  |  |  |  |  |  |  |

Of the subjects included, 1,811 (55.4%) were randomized to WATCHMAN, 1,071 (32.8%) were randomized to Amulet, and 382 (11.7%) were randomized to OACs (N=3,264 subjects overall). Average treatment age for each intervention ranged from 71.7 to 76.5 years, and average CHADSVASC score ranged from 2.0-2.8,^21^ while the average HAS-BLED^22^ score (among the 3 head-to-head trials) ranged from 3.1-4.1. Follow-up time ranged from 45 days^18^ to 18 months.^6,8,17^ Full descriptive statistics can be found on eTable 2 in the ***Online Supplement***.

**Table 2:** Efficacy NMA League Table. Relative Risks of the Primary Efficacy Endpoint by Treatment Group

| Table 2. Relative Risks of the Primary Efficacy Endpoint by Treatment Group |  |  |  |  |  |  |
| --- | --- | --- | --- | --- | --- | --- |
|  | Risk of Stroke: RR [95% CI] |  |  | Risk of Death: RR [95% CI] |  |  |
|  | vs Amulet | vs Watchman | vs OAC | vs Amulet | vs Watchman | vs OAC |
| Amulet |  | 0.95 [0.50; 1.81] | 0.86 [0.35; 2.14] |  | 0.75 [0.55; 1.02] | 0.53 [0.33; 0.85] |
| Watchman |  |  | 1.11 [0.58; 2.10] |  |  | 1.42 [0.98; 2.06] |
| OAC |  |  |  |  |  |  |

### Efficacy Outcomes

All 5 eligible trials reported risk of any stroke and were included in this analysis. Total event counts and forest plots for all outcomes can be found in the ***Online Supplement***. The primary efficacy outcome results are displayed in **Table 2**. The observed risk of any stroke, when compared to OAC, was non-significantly lower in subjects randomized to WATCHMAN (RR=0.90, 95% CI: 0.48, 1.72) and Amulet (RR=0.86, 95% CI: 0.35, 2.14). Head-to-head device comparison suggested no difference in risk of stroke with Amulet (RR=0.95, 95% CI: 0.50, 1.81). The overall I^2^ was 13.4% (Cochran’s Q p=0.33), indicating low risk of heterogeneity bias. Funnel plots did not show any meaningful deviation across trials (***Supplement***). Likewise, all 5 trials reported all-cause death and were included. The risk of death from any cause, however, significantly favored Amulet (RR=0.53, 95% CI: 0.33, 0.85) and non-significantly favored WATCHMAN (RR=0.70, 95% CI: 0.49, 1.02) over OAC. Head to head, Amulet was associated with borderline-significant lower risk of death than WATCHMAN (RR=0.74, 95% CI: 0.55, 1.02, p=0.06). The P-score for Amulet was 0.982, signifying a 98% probability that it is superior in risk of death versus alternative interventions, whereas the probability that OACs were inferior to both alternatives was approximately 98% (Inverse P-score 0.98). The I^2^ estimate was 0% (Cochran’s Q p=0.75), and funnel plots were unremarkable (***Supplement***).

### Safety Outcomes

#### Thromboembolism

The safety outcome results are displayed in **Table 3**. Four of the 5 trials reported risk of any thromboembolic event and were included. When compared to OACs, risk of any thromboembolism was non-significantly increased in both WATCHMAN (RR=2.04, 95% CI: 0.23, 18.4) and Amulet (RR=1.54, 95% CI: 0.11, 22.1). Head-to-head comparison suggested non-significantly lower risk of thromboembolism with Amulet (RR=0.76, 95% CI: 0.17, 3.38). Risk of heterogeneity bias was estimated to be low (I^2^=0%, Cochran’s Q p=0.95). Funnel plots were again unrevealing (***Supplement***).

**Table 3:** Safety NMA League Table. Relative Risks of the Safety Outcomes by Treatment Group

| Table 3. Relative Risks of the Safety Outcomes by Treatment Group |  |  |  |  |  |  |  |  |  |
| --- | --- | --- | --- | --- | --- | --- | --- | --- | --- |
|  | Risk of Thromboembolism: RR [95% CI] |  |  | Risk of Device Embolization: RR [95% CI] |  |  | Risk of Pericardial Effusion: RR [95% CI] |  |  |
|  | vs Amulet | vs Watchman | vs OAC | vs Amulet | vs Watchman | vs OAC | vs Amulet | vs Watchman | vs OAC |
| Amulet |  | 0.76 [0.17; 3.38] | 1.54 [0.11; 22.1] |  | 2.38 [0.67; 8.43] | 7.35 [0.62; 86.6] |  | 2.11 [1.41; 3.17] | 27.0 [3.48; 210] |
| Watchman |  |  | 0.49 [0.054; 4.41] |  |  | 3.10 [0.37; 25.6] |  |  | 12.8 [1.72; 95.2] |
| OAC |  |  |  |  |  |  |  |  |  |

#### Device Embolization

Because risk of device embolization is non-existent with OACs, we report only head-to-head risk ratios for this outcome. All five trials reported this event and were analyzed. Risk of device embolization was non-significantly higher with Amulet when compared to WATCHMAN (RR=2.38, 95% CI: 0.67, 8.43). Estimated risk of heterogeneity bias was low (I^2^=0%, Cochran’s Q p=0.91). Funnel plots were again unremarkable (***Supplement***).

#### Pericardial Effusion (with or without tamponade)

Pericardial effusion, with or without tamponade, was reported in 4/5 trials. Trials varied in their reporting of these effusions. For example, some subcategorized effusions as ‘serious’ versus ‘non-serious’, ‘with tamponade’ or ‘without tamponade’, and ‘treatment-related’ or ‘treatment-unrelated’. Here, we report all cumulative event counts for each group.

When compared to OAC, risk of any pericardial effusion was, as expected, significantly greater with Amulet (RR=27.0, 95% CI: 3.48, 210) and significantly greater with WATCHMAN (RR=12.8, 95% CI: 1.72, 95.3). Head-to-head, risk of effusion over two-fold greater with Amulet than WATCHMAN (RR=2.11, 95% CI: 1.41, 3.17). The P-score suggested that the probability of increased risk with Amulet compared to alternatives was greater than 99% (Inverse P-score=0.9995), and the probability that OACs carried less risk was likewise greater than 99% (P-score=0.996). Risk of heterogeneity bias was again estimated to be low (I^2^=6.3%, Cochran’s Q p=0.34), and funnel plots revealed no meaningful deviations across trials (***Supplement***).

#### Sensitivity Analysis

Because there was concern for bias related to poor relative performance of implant during the initial non-inferiority trials (i.e., versus OACs), a sensitivity analysis was performed to assess whether interpretation might change when excluding these trials. The ***Online Supplement*** displays the forest plots and heterogeneity assessments for each outcome when excluding these two original trials. In brief, estimates were similar to main analysis. However, analyses were possibly underpowered. For example, risk of death with Amulet was 25% lower than with Watchman (RR=0.75, 95% CI: 0.55, 1.02).

## Discussion

To our knowledge, this is the most comprehensive analysis of available RCT evidence on efficacy and safety of LAAC devices. While both Watchman and Amulet serve the purpose of occluding the LAA, differences in design lead to differences in deployment protocol and subsequent efficacy and safety. Previous meta-analyses were either underpowered (lacking utilization of indirect evidence) or more confounded (included cohort studies).

Results from the PROTECT-AF,^8^ PREVAIL,^17^ and long-term follow up of the PROTECT-AF^23^ trials showed that the Watchman device was non-inferior to warfarin in reducing risk of stroke in patients with atrial fibrillation. In the PROTECT-AF trial, Watchman was non-inferior to warfarin therapy in reducing stroke, systemic embolism, and cardiovascular death (RR 0.62, 95% CI 0.35-1.25) with non-inferiority probability being greater than 99.9%. The PREVAIL trial also showed non-inferiority of Watchman compared to warfarin regarding risk of ischemic stroke and systemic embolism (RR 1.60, 95% CI 0.50-4.20).^17^ Furthermore, safety events appeared to occur less frequently in the 7 days following device implant in PREVAIL compared to PROTECT-AF (4.2% vs 8.7%, P=0.004), suggesting improved operator performance as device familiarity increased.

Results from the Amulet IDE suggest that rates of successful deployment – defined as no significant leak post-procedure – are approximately 2.1% higher with Amulet compared to Watchman.^6^ This RCT also demonstrated that the Amulet was noninferior to Watchman for prevention of stroke and systemic embolism.^6^

In this study, we use NMA to increase the statistical power to detect differences between interventions for underpowered secondary outcomes.^15^ Given that death was an underpowered ‘safety’ endpoint (though often included in efficacy composite endpoints) in these studies, it is unsurprising that associations were non-significant in individual trials. Our results suggest a decreased risk of death with Amulet which was statistically significant (RR=0.53, 95% CI: 0.33, 0.85) compared to OAC and bordered significance compared to Watchman (RR=0.74, 95% CI: .55, 1.02). The Amulet’s P-score for death, or probability that it is superior to all other interventions, exceeded 99%. While the mechanism for this difference is not possible to confirm with study-level meta-analysis, it is likely multifactorial. It may partially be attributable to the greater success rate of LAA seal – and discontinuation of anticoagulation – in Amulet recipients.^6,18,19^ Future works using patient-level data should confirm and expand these findings with multivariable regression to rule out confounding.

Although the Amulet IDE demonstrated higher rates of successful deployment and similar efficacy with Amulet compared to Watchman, procedural complications-most significantly pericardial effusion and device embolization-were more likely to occur with Amulet (4.5% versus 2.5%).^6^ Our NMA demonstrated a two-fold greater risk of effusion with Amulet than WATCHMAN (RR=2.11, 95% CI: 1.41, 3.17). Operators may presently prefer WATCHMAN over Amulet due to this increased risk of late pericardial effusion, which can result in life-threatening cardiac tamponade.^6^ Indeed, risk of any pericardial effusion, with or without tamponade, was more than two-fold greater with Amulet than WATCHMAN in this analysis. The Amulet, once deployed, typically has greater surface contact with the LAA which subsequently results in inflammation and effusion.^6^ The risk is exacerbated by operator inexperience with deploying Amulet, and notably no effusions occurred in subjects undergoing implant with an interventionalist with >9 cases of experience.^6^

## Limitations

This study is not without limitations. As previously noted, NMAs use all available evidence to generate effect estimates for each intervention. Because these estimates are therefore non-randomized, we refrain from interpreting these results causally. This is especially true for Amulet versus OAC comparisons, which are based exclusively on indirect evidence. Previous direct meta-analysis^24^ compared only WATCHMAN and Amulet but 16/19 studies included were observational. Thus, it was more subject to bias, yet also did not include indirect RCT evidence and thereby lacked statistical power to properly evaluate outcomes such as death.

The only OAC included in this study is warfarin, which is highly effective in preventing AF-related ischemic stroke^25^ but has an unfavorable safety profile when compared with Factor Xa inhibitors such as apixaban.^26^ This limits interpretation of comparative efficacy and safety of OACs, but does not impact estimates of device comparison. Our study also combined multiple generations of Watchman devices for comparison with the Amulet LAAC device and OACs. Newer generations of both Amulet and Watchman devices have since been released, which may differ in safety and efficacy when compared to previous versions. However, one included study^18^ indicated differences between Watchman 2.5 and FLX were likely incremental and therefore might reasonably combined. Further subcategorization by generation would be preferable, but there is insufficient power for such analyses. Exclusion of the two earliest trials (Watchman vs. OAC) did not change the estimate for death risk in Amulet versus Watchman.

Study-level differences in 1) exclusion criteria, 2) operator experience, and 3) event definitions could have meaningfully changed event rates and confounded our findings. For example, the exclusion of subjects with a prior history of major bleeding in the studies where subjects were randomized to OACs or WATCHMAN^8,17^ almost certainly decreased absolute risk of death and hemorrhagic stroke within the trial, while inclusion of inexperienced operators in the Amulet-IDE trial likely increased risk of pericardial effusion.^27^ While the first issue might limit power in detecting underlying effects, it would not change relative risk estimates. NMAs overcome this limitation by utilizing all available evidence, whereas traditional meta-analyses could have failed to detect the differences described here. The second issue suggests that adverse event rates attributable to the procedure in these trials are most generalizable in settings where practitioners have little experience with LAA closure. The issue of trial differences in event definitions was mitigated by our decision to not distinguish, for example, between ‘treatment-related’ vs. ‘treatment-unrelated’ events and other event subclassifications. While this approach protects against confounding from lack of blinding and/or definitional differences across studies, it provides less precise information on the seriousness of these events.

Further, our findings are consistent with recent real-world data,^27^ were estimated to be at low risk for heterogeneity bias, and did not appear affected by any outlier results. The findings are substantial enough to warrant individual patient meta-analysis and possibly larger-scale, long-term head-to-head trials between newer generations of Watchman and novel editions of Amulet.^24^ An individual patient data meta-analysis is the best method for ameliorating these limitations, and the logical next step in evaluating these devices’ comparative efficacy and safety. Abstracting study-level data can also lead to miscoding or misinterpretation of reported outcomes. We minimized this risk by having multiple study members code each article blindly, and strictly reporting trial-definition/reported outcomes rather than imposing a universal event definition. We minimized risk of confounding by qualitatively assessing funnel plots for skew, and quantitatively assessing risk of heterogeneity bias across trials. While these methods cannot rule out confounding, they strongly discourage the possibility that results would change with multivariable regression. The small number of trials precluded formal hypothesis tests for assessing bias, but given the low estimated risk of heterogeneity bias it is unlikely this limitation could impact confidence or interpretation of these findings.

## Conclusion

This study suggests a meaningful mortality benefit with Amulet compared to both WATCHMAN and OACs, despite a large increased risk of perioperative pericardial effusion in inexperienced operators. Pooled patient analysis of RCTs is indicated to further elucidate this relationship.

## Data Availability

Data will be shared on reasonable request. Analyses, including code for R, are available at rpubs.com for reproducibility. This page is live and may be supplemented after study publication.

