## Supplementary material for "COMPARING EFFICACY AND SAFETY OF LEFT ATRIAL APPENDAGE CLOSURE DEVICES: A NETWORK META-ANALYSIS OF RANDOMIZED CONTROLLED TRIALS": Figure 1 (from another version of this preprint: version 2, 2024-04-01)

**5 unique, eligible trials identified**

Articles excluded for:

**Wrong Study Design** (n = 5)

**Mixed Protocol** (i.e., combined LAAC Devices vs OAC) (n = 5)

**No Eligible Outcomes** (n = 18)

Articles Excluded:

**Wrong Study Design, Mixed Interventions, and/or No Relevant Outcomes Data**

(n = 399)

Articles Excluded:

**Duplicates**

(n = 9)

**Full-text articles assess for eligibility**

(n = 33)

18 Sys. Reviews, Meta-Analyses

13 RCTs

**Total records screened by title and abstract**

(n = 432)

**Records identified through PubMed, Cochrane, Web of Science Databases**

(RCTs, Systematic Reviews, Meta-Analyses)

(n = 441)
