## Supplementary material for "COMPARING EFFICACY AND SAFETY OF LEFT ATRIAL APPENDAGE CLOSURE DEVICES: A NETWORK META-ANALYSIS OF RANDOMIZED CONTROLLED TRIALS": Online Supplement (from another version of this preprint: version 2, 2024-04-01)

### Search Strategy (Ovid only)

```
Ovid Technologies, Inc. Email Service

------------------------------

Search for: 38 not 39

Results: 337

Database: Ovid MEDLINE(R) and Epub Ahead of Print, In-Process, In-Data-Review & Other Non-Indexed Citations and Daily
<1946 to May 30, 2023>
Search Strategy:
--------------------------------------------------------------------------------
1     exp Atrial Fibrillation/ (70532)
2     (atrial adj5 fibrillat*).mp. (104814)
3     (auricular* adj5 fibrillat*).mp. (1591)
4     1 or 2 or 3 (104978)
5     exp Septal Occluder Device/ (3492)
6     amplatzer*.mp. (3353)
7     cardioseal*.mp. (122)
8     septal occluder*.mp. (4216)
9     5 or 6 or 7 or 8 (5842)
10     4 and 9 (935)
11     watchman*.mp. (855)
12     (left atrial adj3 occlusion*).mp. (1409)
13     (left atrial adj3 occlud*).mp. (191)
14     (left atrium adj3 occlusion*).mp. (24)
15     (left atrium adj3 occlud*).mp. (20)
16     (left atrial adj3 closure*).mp. (1573)
17     (left atrium adj3 closure*).mp. (35)
18     11 or 12 or 13 or 14 or 15 or 16 or 17 (3051)
19     4 and 18 (2515)
20     10 or 19 (2772)
21     limit 20 to english language (2639)
22     limit 21 to (systematic reviews pre 2019 or systematic reviews) (175)
23     limit 21 to (meta analysis or "systematic review") (119)
24     22 or 23 (181)
25     limit 24 to yr="2020 -Current" (85)
26     limit 21 to randomized controlled trial (55)
27     exp Placebos/ (39461)
28     random$.tw. (1419026)
29     double blind$.tw. (169904)
30     double dummy.tw. (2471)
31     mask$.tw. (99814)
32     sham.tw. (99861)
33     placebo$.tw. (246272)
34     (control$ adj trial$).mp. (896702)
35     exp Randomized Controlled Trial/ (595179)
36     27 or 28 or 29 or 30 or 31 or 32 or 33 or 34 or 35 (1863754)
37     21 and 36 (356)
38     24 or 37 (441)
39     9 and 18 and 38 (104)
40     38 not 39 (337)

***************************
```

### Trial Information and Data Analysis Coding

| **eTable 2: Descriptives and Outcomes by Trial** | | | | | | | | | | | | | | | | | | | | | | | | |
| --- | --- | --- | --- | --- | --- | --- | --- | --- | --- | --- | --- | --- | --- | --- | --- | --- | --- | --- | --- | --- | --- | --- | --- | --- |
| **Study Details** | | | | | | | | **Descriptives and Comorbid Conditions** | | | | | | | | | **Outcomes** | | | | | | | |
| **Author, Year, Trial Name** | **Intervention 1** | **Intervention 2** | **Sample 1** | **Sample 2** | **History of HTN 1** | **History of HTN 2** | **History of HLD 1** | | **History of HLD 2** | **CHADSVASC2 Score 1** | **CHADSVASC2 Score 2** | **HASBLED Score 1** | **HASBLED Score 2** | **Strokes 1** | **Strokes 2** | **Death 1** | | **Death 2** | **Any Thromboembolism 1** | **Any Thromboembolism 2** | **Device Embolization 1** | **Device Embolization 2** | **Pericardial Effusion 1** | **Pericardial Effusion 2** |
| *Galea et al 2022 (SWISS-APERO)* | Watchman | Amulet | 110 | 111 | 90 | 87 | - | | - | 4.4 | 4.2 | 3.2 | 3.1 | 2 | 2 | 2 | | 4 | 1 | 1 | 1 | 1 | 8 | 22 |
| *Lakkireddy et al 2021 (Amulet IDE)* | Watchman | Amulet | 944 | 934 | - | - | - | | - | 4.7 | 4.5 | 3.3 | 3.2 | 29 | 24 | 85 | | 63 | 3 | 2 | 2 | 6 | 11 | 22 |
| *Mansour et al 2021* | Watchman | Amulet | 25 | 26 | 17 | 21 | 15 | | 12 | 3.9 | 3.9 | 4.2 | 4.1 | 4 | 0 | 1 | | 0 | - | - | 1 | 0 | - | - |
| *Holmes et al 2014 (PREVAIL)* | Watchman | Anticoagulation | 269 | 138 | 238 | 134 | - | | - | 3.8 | 3.9 | - | - | 6 | 1 | 37 | | 25 | 1 | 0 | 2 | 0 | 4 | 0 |
| *Holmes et al 2009 (PROTECT AF)* | Watchman | Anticoagulation | 463 | 244 | 413 | 220 | - | | - | 2.0* | 2.3* | - | - | 16 | 12 | 21 | | 18 | 2 | 0 | 3 | 0 | 22 | 0 |
| ** CHADS2 Score Reported* | | | | | | | | | | | | | | | | | | | | | | | | |

### Funnel Plot and Heterogeneity Assessment: Risk of Any Stroke

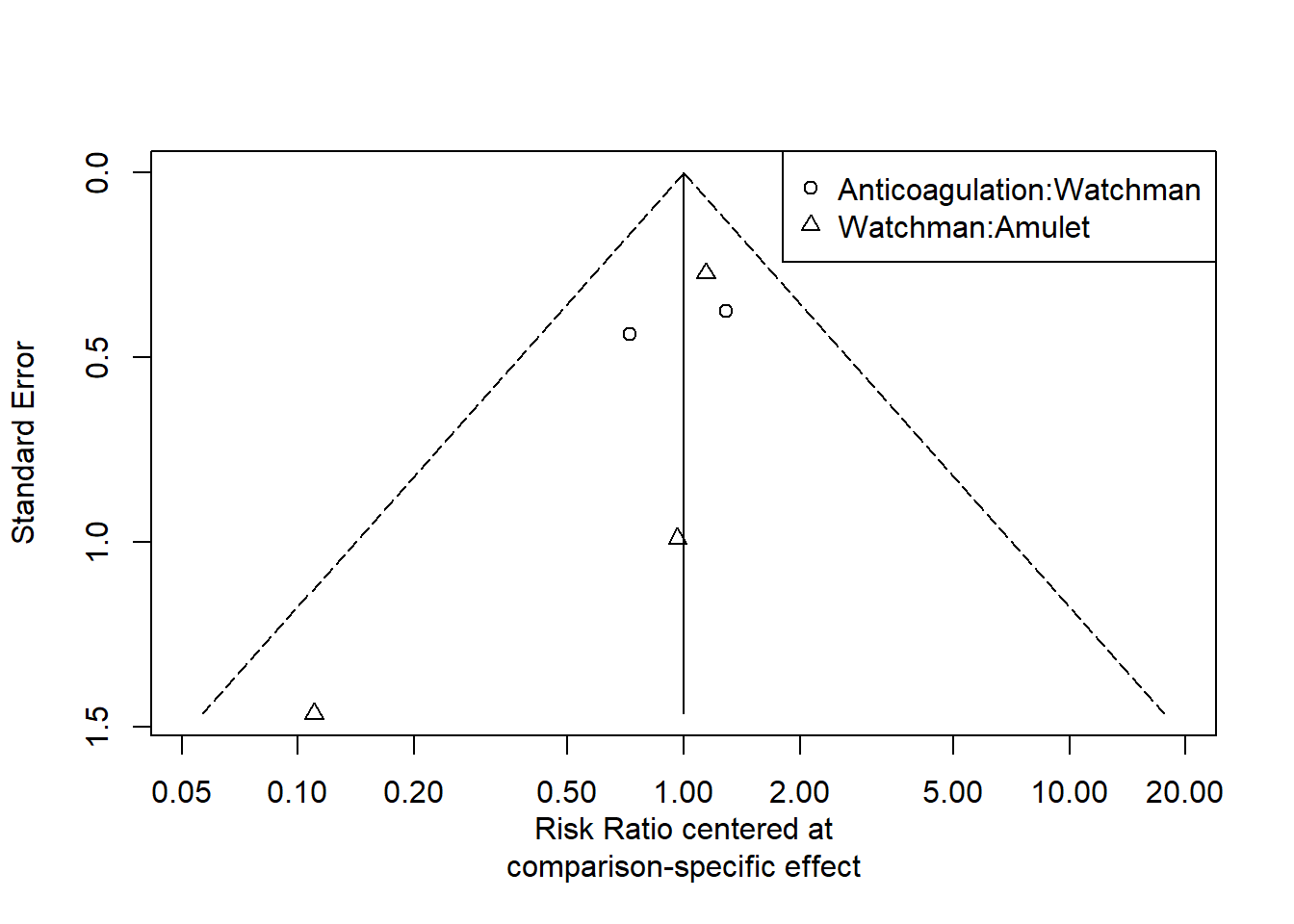

```
## Quantifying heterogeneity / inconsistency:
## tau^2 = 0.0526; tau = 0.2294; I^2 = 13.4% [0.0%; 86.7%]
##
## Tests of heterogeneity (within designs) and inconsistency (between designs):
##                    Q d.f. p-value
## Total           3.46    3  0.3256
## Within designs  3.46    3  0.3256
## Between designs 0.00    0      --
```

### Funnel Plot and Heterogeneity Assessment: Risk of Death (Any Cause)

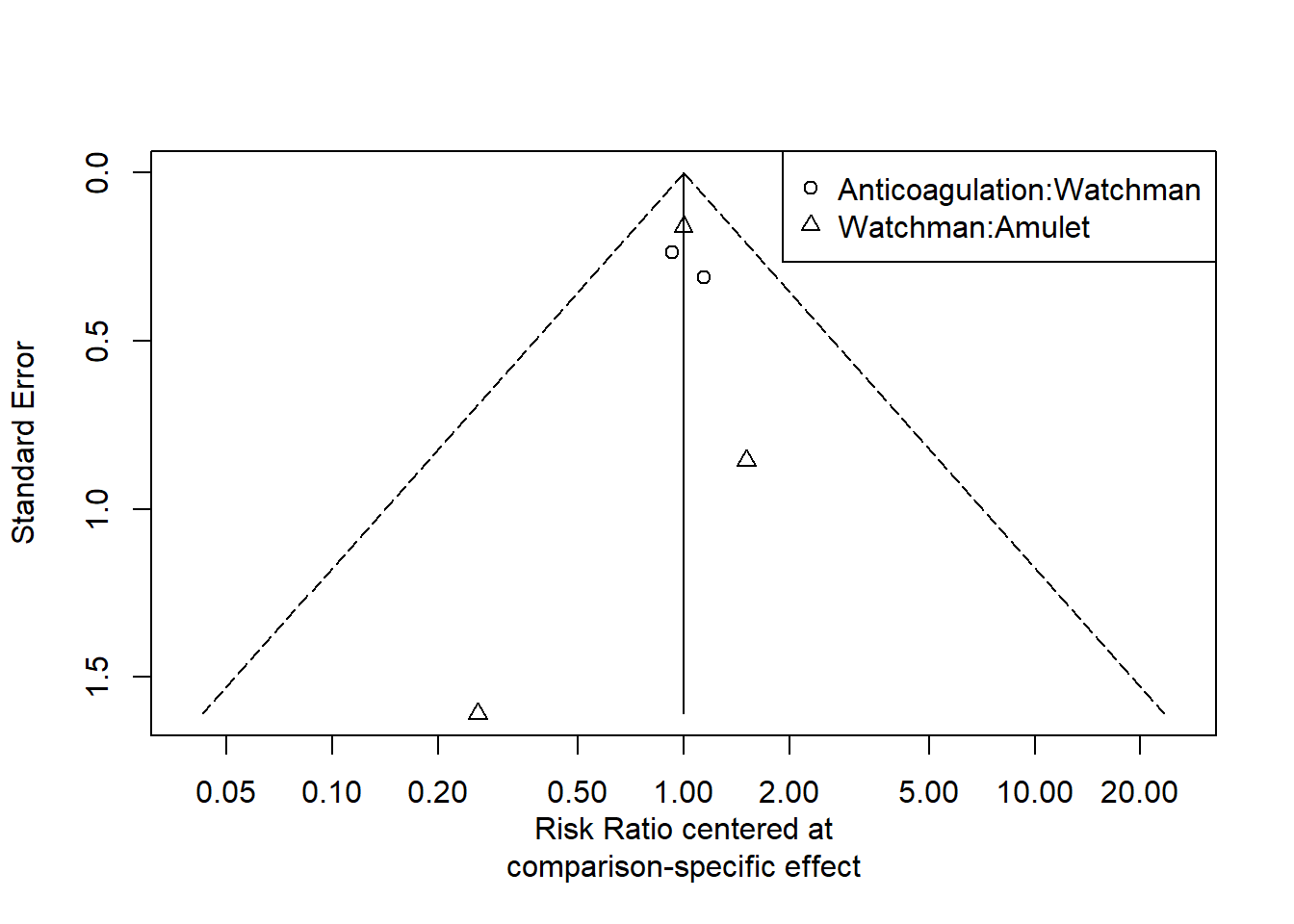

```
## Quantifying heterogeneity / inconsistency:
## tau^2 = 0; tau = 0; I^2 = 0% [0.0%; 84.7%]
##
## Tests of heterogeneity (within designs) and inconsistency (between designs):
##                    Q d.f. p-value
## Total           1.23    3  0.7465
## Within designs  1.23    3  0.7465
## Between designs 0.00    0      --
```

### Funnel Plot and Heterogeneity Assessment: Risk of Thromboembolism

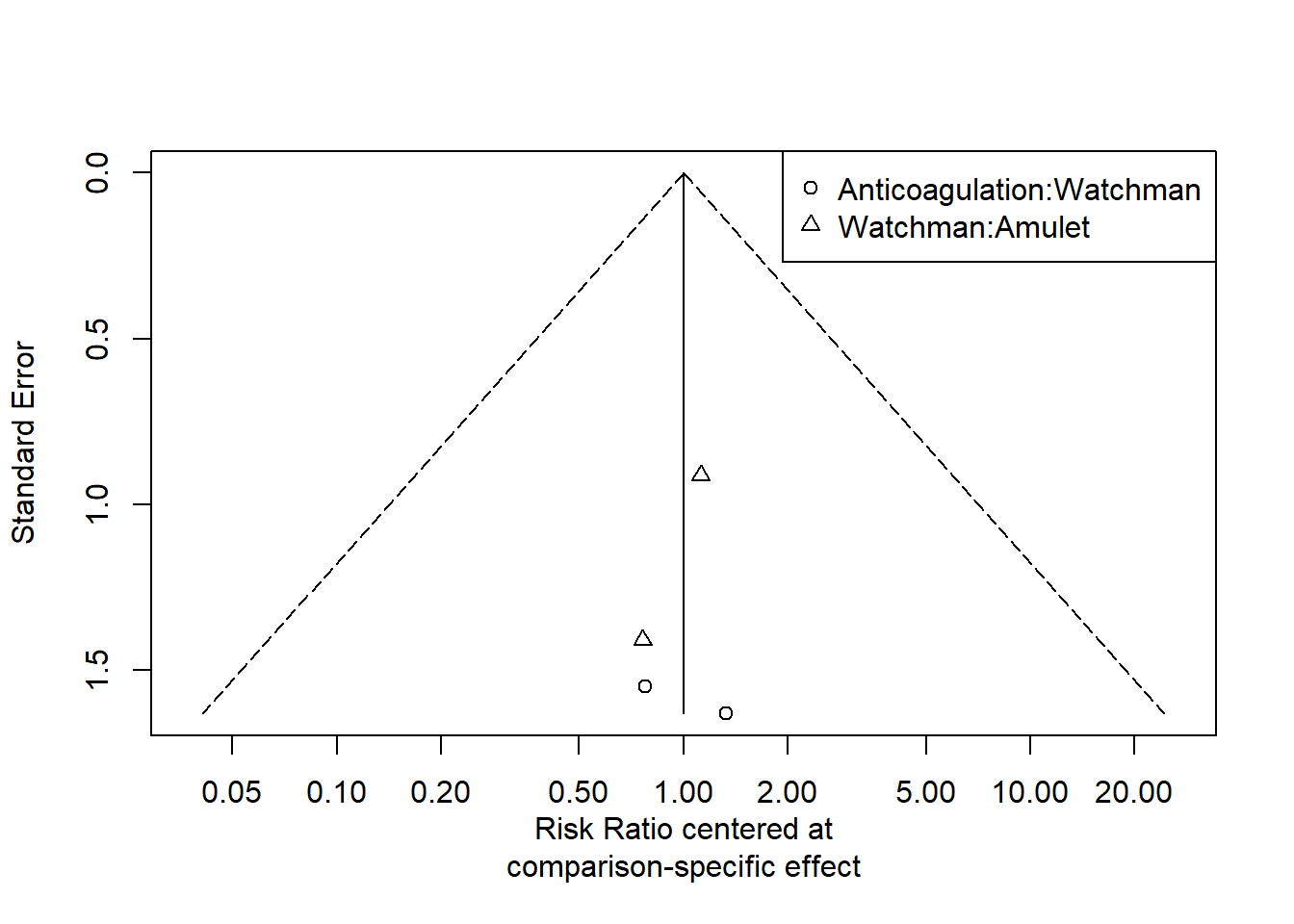

```
## Quantifying heterogeneity / inconsistency:
## tau^2 = 0; tau = 0; I^2 = 0% [0.0%; 89.6%]
##
## Tests of heterogeneity (within designs) and inconsistency (between designs):
##                    Q d.f. p-value
## Total           0.11    2  0.9465
## Within designs  0.11    2  0.9465
## Between designs 0.00    0      --
```

### Funnel Plot and Heterogeneity Assessment: Risk of Device Embolization

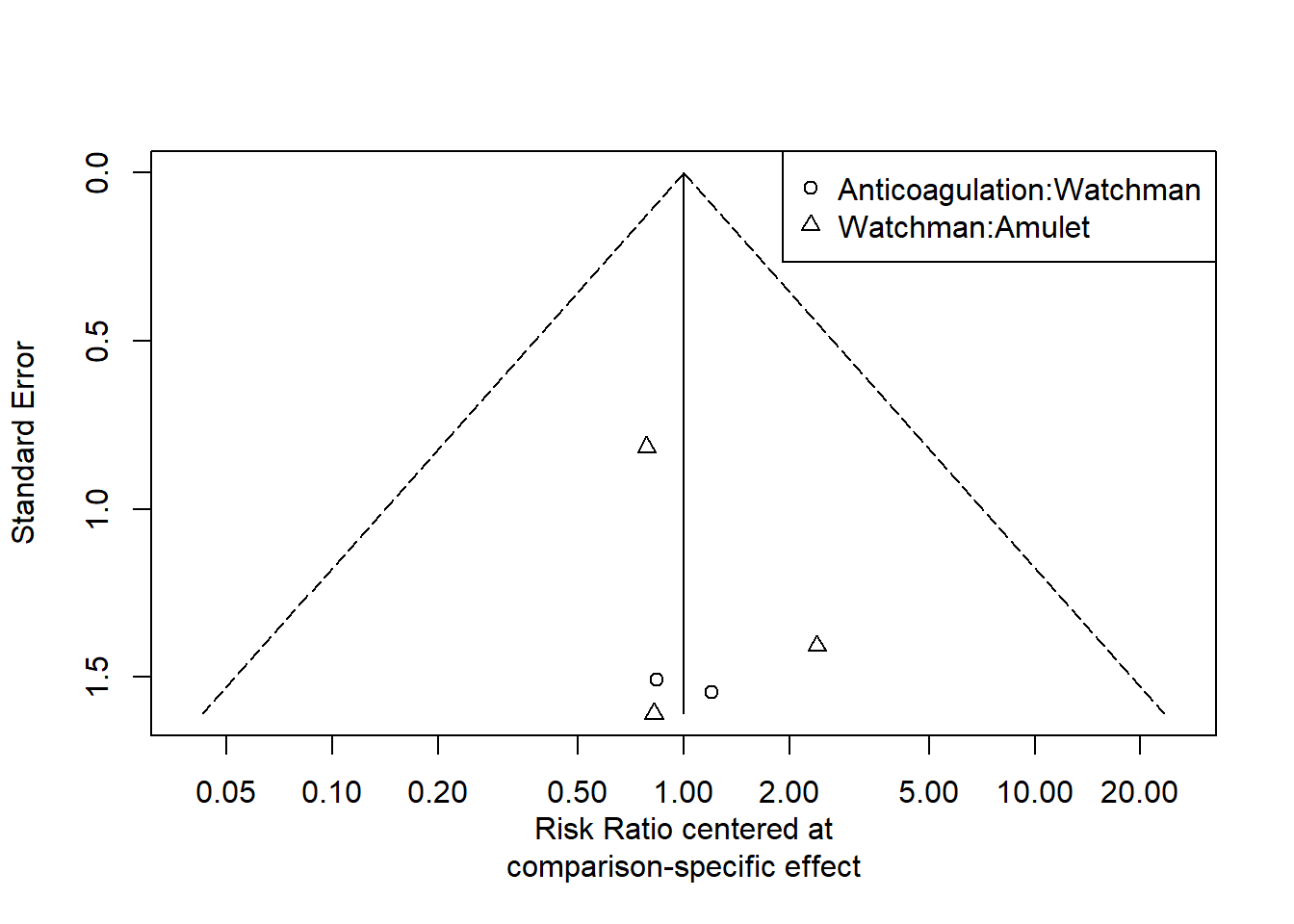

```
## Quantifying heterogeneity / inconsistency:
## tau^2 = 0; tau = 0; I^2 = 0% [0.0%; 84.7%]
##
## Tests of heterogeneity (within designs) and inconsistency (between designs):
##                    Q d.f. p-value
## Total           0.52    3  0.9149
## Within designs  0.52    3  0.9149
## Between designs 0.00    0      --
```

### Funnel Plot and Heterogeneity Assessment: Risk of Pericardial Effusion

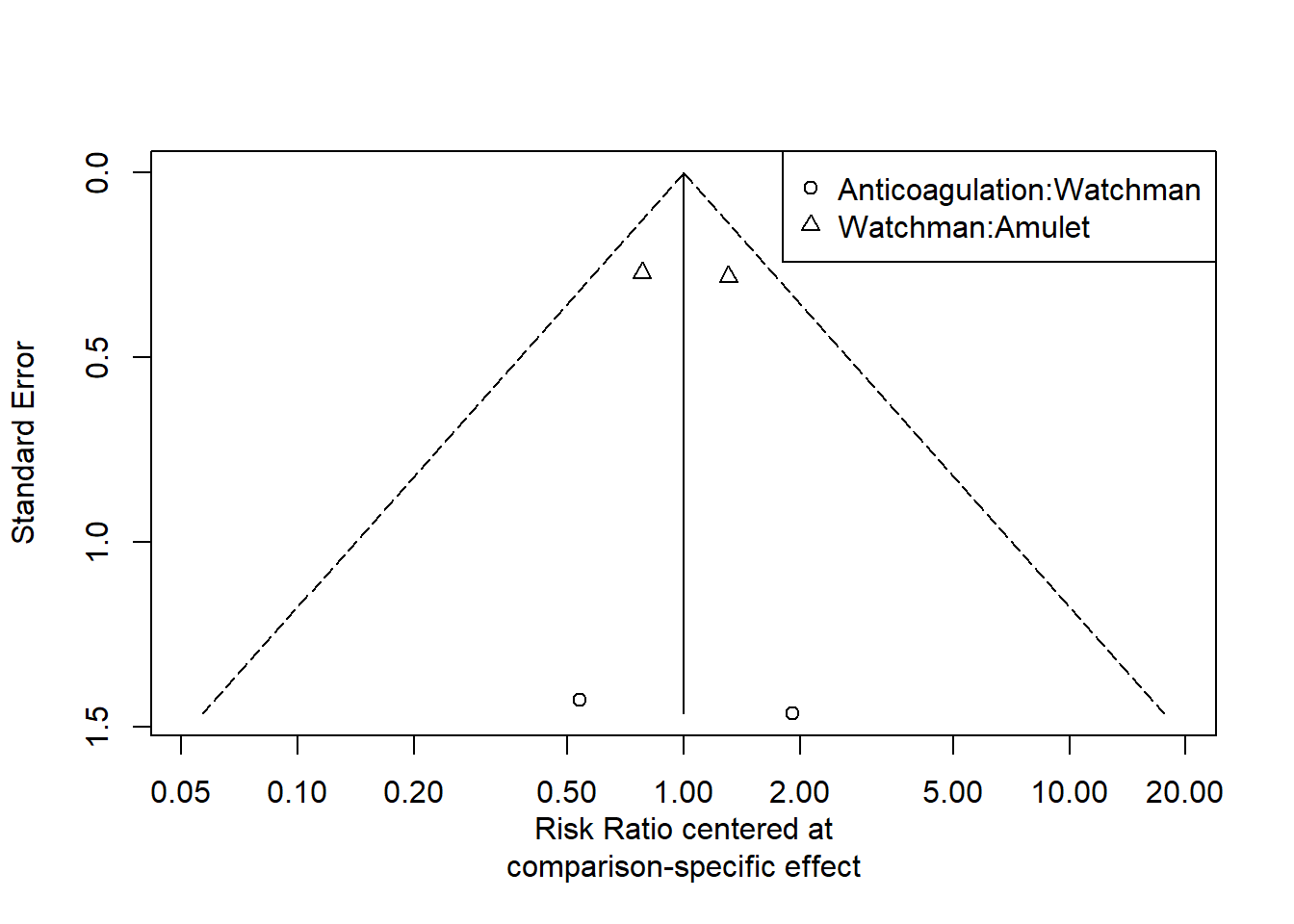

```
## Quantifying heterogeneity / inconsistency:
## tau^2 = 0.0098; tau = 0.0992; I^2 = 6.3% [0.0%; 90.3%]
##
## Tests of heterogeneity (within designs) and inconsistency (between designs):
##                    Q d.f. p-value
## Total           2.13    2  0.3440
## Within designs  2.13    2  0.3440
## Between designs 0.00    0      --
```
